# Strength of Statistical Evidence for the Efficacy of Cancer Drugs: A Bayesian Re-Analysis of Randomized Trials Supporting FDA Approval

**DOI:** 10.1101/2023.06.30.23292074

**Authors:** Merle-Marie Pittelkow, Maximilian Linde, Ymkje Anna de Vries, Lars G. Hemkens, Andreas M. Schmitt, Rob R. Meijer, Don van Ravenzwaaij

## Abstract

**Objective:** To quantify the strength of statistical evidence of randomised controlled trials (RCTs) for novel cancer drugs approved by the Food and Drug Administration (FDA) in the last two decades.

**Study Design and Setting:** We used data on overall survival (OS), progression-free survival (PFS), and tumour response (TR) for novel cancer drugs approved for the first time by the FDA between January 2000 and December 2020. We assessed strength of statistical evidence by calculating Bayes Factors (*BF*s) for all available endpoints, and we pooled evidence using Bayesian fixed-effect meta-analysis for indications approved based on two RCTs. Strength of statistical evidence was compared between endpoints, approval pathways, lines of treatment, and types of cancer.

**Results:** We analysed the available data from 82 RCTs corresponding to 68 indications supported by a single RCT and seven indications supported by two RCTs. Median strength of statistical evidence was ambiguous for OS (*BF* = 1.9; IQR 0.5-14.5), and strong for PFS (*BF* = 24,767.8; IQR 109.0-7.3*10^6^) and TR (*BF* = 113.9; IQR 3.0-547,100). Overall, 44 indications (58.7%) were approved without clear statistical evidence for OS improvements and seven indications (9.3%) were approved without statistical evidence for improvements on any endpoint. Strength of statistical evidence was lower for accelerated approval compared to non-accelerated approval across all three endpoints. No meaningful differences were observed for line of treatment and cancer type.

**Limitations:** This analysis is limited to statistical evidence. We did not consider non-statistical factors (e.g., risk of bias, quality of the evidence).

**Conclusion:** *BF*s offer novel insights into the strength of statistical evidence underlying cancer drug approvals. Most novel cancer drugs lack strong statistical evidence that they improve OS, and a few lack statistical evidence for efficacy altogether. These cases require a transparent and clear explanation. When evidence is ambiguous, additional post-marketing trials could reduce uncertainty.

## BACKGROUND

For a new cancer drug to be marketed in the U.S., it needs to be endorsed by the Food and Drug Administration (FDA). While many aspects of a drug’s profile are considered in the approval process, the statistical evaluation of efficacy plays a central role [1]. Cancer drugs are frequently approved based on limited evidence, which increases uncertainty in clinical decision making [2–7]. Despite FDA guidelines suggesting that substantial evidence for efficacy based on two convincing trials should be provided [8], approval for novel cancer drugs between 2000 and 2020 was typically based on a single pivotal (i.e., efficacy determining) trial [2,7]. The understanding that this leads to a reduced level of statistical evidence for efficacy is implicit. Previous work has considered the strength of evidence indirectly considering effect sizes, number and kinds of trials, or qualitative evidence [2,7].

Explicit quantification of the statistical strength of evidence is missing. Another complication arises from the FDA permitting surrogate endpoints, that is, outcomes that are no “direct measurement[s] of clinical benefit but [] known to predict clinical benefit” (p. 16 [9]) such as overall survival (OS) but are easier or faster to measure [10]. However, evidence that surrogate endpoints, like progression-free survival (PFS) and tumour response (TR), predict overall survival in oncology is limited [10–12], limiting the quality of the evidence available at the time of approval. Questions have been raised whether the current statistical evidence for cancer drugs meets the FDA’s requirement for showing “meaningful therapeutic benefits” [3].

Explicit quantification of statistical strength of evidence for efficacy at the time of approval can be achieved with Bayes Factors (*BF*s). Technically, *BF*s compare the likelihood of the observed data under the null hypothesis (i.e., novel treatment does not improve outcomes) to the likelihood of the observed data under an alternative hypothesis (e.g., novel treatment improves outcomes; one-sided alternative hypothesis) [13–15]. The resulting ratio provides a relative measure of statistical evidence for or against competing hypotheses (see Figure 1). For example, a *BF* of 1 indicates ambiguity as the observed study results are equally likely to have occurred if the novel treatment improves OS or if the novel treatment does not improve OS. A *BF* of 10 indicates that the observed study results are ten times more likely to have occurred given that the novel treatment improves OS than if it does not improve OS, whereas a *BF* of 0.1 indicates that the observed study results are ten times more likely to have occurred given that the novel treatment does not work than if it does work.

**Figure 1.**
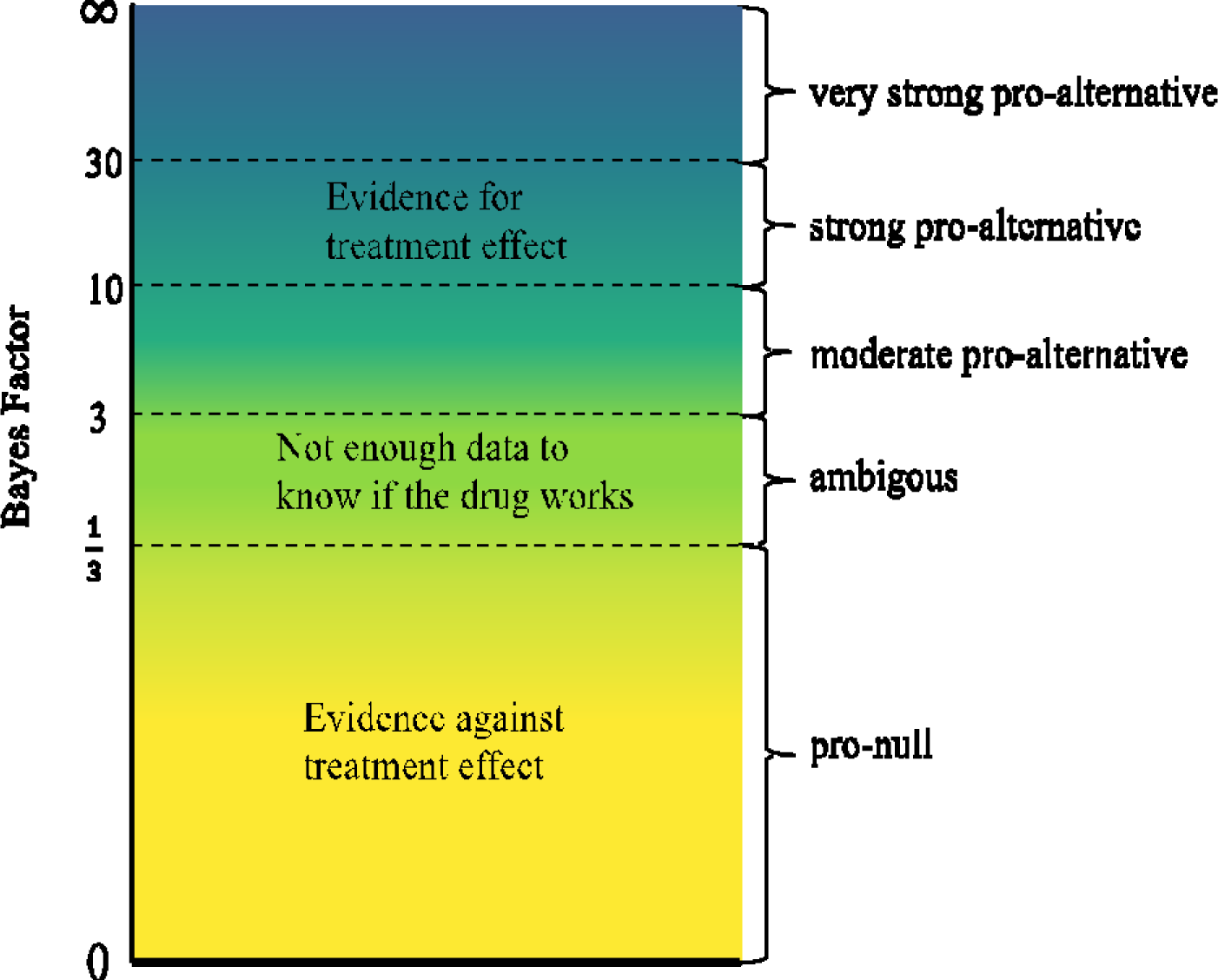
Visual representation of the continuous properties and thresholds of the Bayes Factor in the context of treatment evaluation. Bayes Factors above 3 indicate evidence in favour of the treatment effect. Bayes Factors below 1/3 indicate evidence against the treatment effect. Note: These thresholds have been proposed by methodological researchers and are not clinically informed [15,24].

Using the *BF,* we offer a different perspective on statistical evidence that may be more intuitive than the traditional focus on statistical significance in frequentist frameworks. We take a perspective like that of a clinician who wants to know if the results of a diagnostic test (here, the approval trials) are more likely under a working diagnosis than under a differential diagnosis, which is reflected by the *BF*. Moreover, using *BF*s, unlike traditional Frequentist testing, enables one to differentiate between absence of evidence (i.e., ambiguous evidence) and evidence of absence (i.e., pro-null evidence). This project has two aims. First, we aim to quantify and describe the strength of statistical evidence for efficacy associated with novel^1^ cancer drugs approved between 2000 and 2020 using *BF*s. Our second aim is to contrast strength of statistical evidence for efficacy between endpoints, approval types, lines of treatment, and type of cancer, as standards for what constitutes “sufficient” strength of statistical evidence for beneficial effects (and against harmful effects) at the time of approval might differ. For example, one might accept more uncertainty and lower strength of statistical evidence for 3rd or 4th line treatments than for 1st or 2^nd^ line treatments. Previous reports also suggest that novel drugs for haematological cancers are more likely to be approved based on single-arm trials and surrogate endpoints compared to solid cancers [2].

## METHOD

### Data and Registration

The aim and general approach of the project were registered at OSF (https://osf.io/exyfd/). Analysis code and data are available from https://ceit-cancer.org/ and OSF (https://osf.io/4uhz7). All files needed to reproduce the analyses are available on OSF (https://osf.io/qz7xy/).

### FDA Data

We used data from the CEIT-Cancer project (details provided elsewhere [16,17]). In short, novel drugs and biological therapies receiving first approval for the treatment of any malignant diseases between January 2000 and December 2020 and the corresponding FDA reviews (available at <u>drugs@FDA</u>) were identified. For all RCTs evaluating the drug in the approved indication (regardless of whether the trial was described as pivotal or not), the following data were extracted: hazard ratios (HR) and associated 95% confidence intervals for OS and/or PFS; median OS and/or PFS; number of events for OS, PFS and/or TR; sample size; line of treatment; approval pathway (i.e., priority review, orphan design, accelerated approval, and breakthrough therapy designation); type of cancer; type of control; and type of blinding. The original data set also included single-arm clinical trials that were explicitly described as pivotal. However, here we only considered data from RCTs as single-arm trials do not provide comparative treatment outcomes.

### Data Analysis

We used R version 4.1.2 [18]. We calculated *BF*s for all available endpoints per RCT. For OS and PFS, we used the available summary statistics (i.e, HR, confidence interval of HR, sample size, and number of events per group) to conduct a Bayesian Cox regression using the “baymedr” R package [19]. We used a standard normal distribution that was truncated at 0 as the prior for beta under the alternative hypothesis. For TR outcomes, *BF*s for chi-square tests were calculated using the “BayesFactor” R package [20]. Under the null hypothesis, the prior for the proportion of TR was a joint uniform distribution ranging from 0 to 1. Under the alternative hypothesis, the prior for the proportion of TR was a uniform distribution ranging from 0 to 1 for each group independently [21]. See supplement (section 1) for further details. For approvals based on two RCTs, we pooled available outcomes for OS and PFS via fixed-effect meta-analysis using the “metaBMA” R package [22]. Under the alternative hypothesis, the prior for the treatment effect was a default Cauchy distribution that was truncated at, with a location parameter of 0 and a scale parameter of 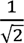 [23]. For TR, we pooled the number of events and number of participants and calculated corresponding chi-square tests using the “BayesFactor” R package with the same specifications as for single trial results. To describe the strength of statistical evidence we adopted standard thresholds (see Figure 1).

The interpretation of pro-null *BF*s depends on the control group. Trials were classified as inactively controlled if the control group received a placebo or no treatment even if all study patients received treatment as usual (e.g., chemotherapy). Trials were classified as actively controlled if the control group received another active treatment different from the experimental group (e.g., an established treatment). In trials with inactive control groups, a pro-null *BF* means that there is evidence that the drug performs comparable to or worse than placebo or no treatment, indicating no efficacy of the novel drug. In trials with an active control group, interpreting pro-null *BF*s is more difficult. Assuming that the active control group receives an effective therapy, a pro-null *BF* can mean that the novel drug is ineffective, or just that it is not *more* effective than an established effective treatment. Therefore, we discuss evidence per control condition whenever possible. For the subgroup analysis of approval pathway, line of treatment, and type of cancer, we did not separate results by control group as these subgroups were too small to be split further. We describe results separately for drugs supported by a single RCT versus drugs supported by two RCTs, as the two RCTs supporting a single drug may use different control groups (i.e., one using an inactive control group and one using an active control group). This decision was not pre-registered.

### Exploratory data analysis

We conducted four not pre-registered analyses, exploring: (1) the relationship between *BFs* and effect sizes, (2) the relationship between BFs and sample size, and (3) the qualitative reasoning behind endorsement decisions approved based on pro-null or ambiguous statistical evidence. Rationales and details are provided in the supplement (section 3), and (4) in response to a reviewer comment we examined the median strength of statistical evidence for primary endpoints and (5) the relationship between evidential strength of OS and PFS, which is reported in the supplement.

## RESULTS

The dataset contained data on 145 novel cancer drugs for 156 indications based on unique 186 trials. Of these, 75 indications (48.1%) received FDA approval without supporting evidence from RCTs, 70 indications (44.9%) were supported by one, and 11 (7.1%) indications were supported by two RCTs. Summary data were unavailable for 10 RCTs. Consequently, we analysed 82 RCTs supporting approval for 75 indications of 75 novel cancer drugs, 68 (90.7%) supported by a single RCT and 7 (8.5%) supported by two RCTs (see Figure S1). Of the 82 included trials, 72 trials assessed OS (87.8%), 66 trials assessed PFS (80.5%), and 68 trials assessed TR (82.9%). Overall, 52 trials provided data for all three endpoints (63.4%), 20 trials data for two endpoints (24.4%), and 10 trials for only one endpoint (12.2%). Trial characteristics and individual *BF*s per trial and endpoint are presented in the supplement (Table S1 and S2).

### Approval decisions based on one RCT

Median evidential strength for drugs approved with one supportive RCT is provided in Table 1, and the distribution of *BF*s per endpoint and control group are presented in Figure 2.

**Figure 2.**
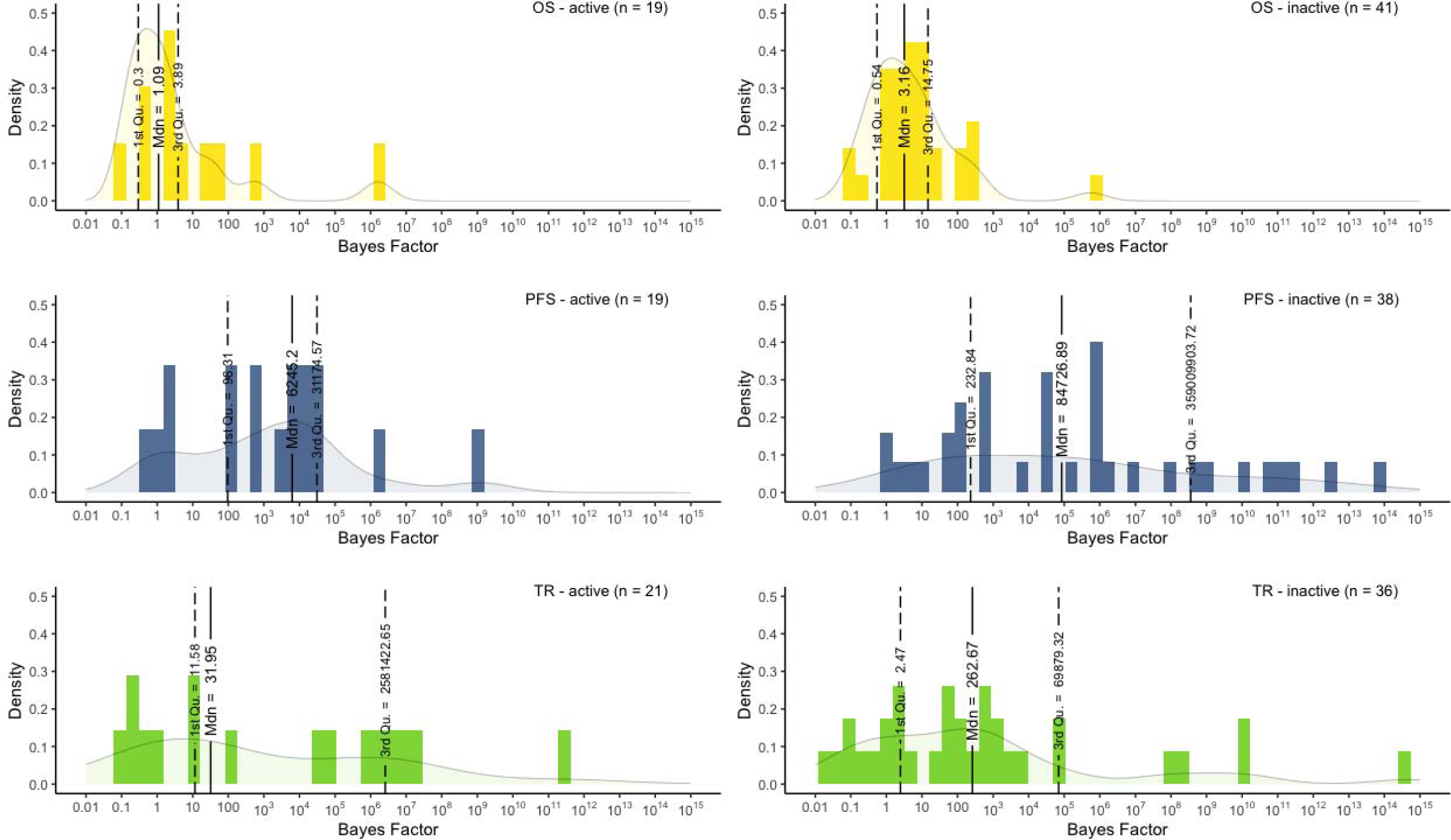
Histograms and density plots illustrating the distribution of BFs for the three possible endpoints and two types of comparators. Medians and first and third quartiles are presented in vertical lines. Note that for PFS-active 2 effects, for PFS-inactive 2 effects, TR-active 1 effect, and TR-inactice 3 effects were cut off.

**Table 1.** Descriptives for drug approvals supported by one RCT.

|  | *Overall* |  | *OS* |  | *PFS* |  | *TR* |  |
| --- | --- | --- | --- | --- | --- | --- | --- | --- |
|  | *N (%)* | *Median BF (IQR)* | *N (%)* | *Median BF [IQR]* | *N (%)* | *Median BF [IQR]* | *N (%)* | *Median BF [IQR]* |
| **Overall** | 68 (100) |  | 60 (100) | 1.9 [0.5; 14.5] | 57 (100) | 24767.8 [109; 7287000] | 57 (83.8) | 114 [3; 547100] |
| **Control group** |  |  |  |  |  |  |  |  |
| active | 22 (32.4) | 17.87 [1.0; 12220] | 19 (31.7) | 1.1 [0.3; 3.9] | 19 (33.3) | 245.2 [96.0; 31180] | 21 (36.8) | 32.0 [12.0; 2581000] |
| inactive | 46 (67.6) | 58.18 [2.0; 47080] | 41 (68.3) | 3.2 [0.5; 14.7] | 38 (66.7) | 84726.9 [233.0; 3.5*10<sup>8</sup>] | 36 (63.2) | 263 [2.0; 69880] |
| **Accelerated approval** |  |  |  |  |  |  |  |  |
| yes | 10 (14.7) | 11.58 [0; 134] | 7 (11.7) | 0.5 [0.3; 16.9] | 7 (12.3) | 426.5 [52.0; 1165904] | 7 (12.3) | 32.0 [12.0; 2581000] |
| no | 58 (85.3) | 41.09 [1.0; 34220] | 53 (88.3) | 2.0 [0.8; 14.4] | 50 (87.7) | 26629.3 [233.0; 3.5*10<sup>8</sup>] | 50 (87.7) | 262.7 [2.0; 69880] |
| **Line of treatment** |  |  |  |  |  |  |  |  |
| first | 31 (45.6) | 17.9 [2.0; 28900] | 30 (50.0) | 2.0 [0.9; 13.0] | 23 (40.4) | 28490.7 [105.0; 5.8 *10<sup>8</sup>] | 24 (42.1) | 621.5 [9.0; 5366000] |
| second | 28 (41.2) | 83.6 [1.0; 64850] | 22 (36.7) | 1.7 [0.5; 13.9] | 26 (45.6) | 2631.4 [418.0; 6060000] | 26 (45.6) | 123.9 [9.0; 435200] |
| ≥ third | 9 (13.2) | 5.00 [0.0; 4310] | 8 (13.3) | 1.7 [0.4; 12.1] | 8 (14.0) | 17761.5 [43.0; 2.1*10<sup>9</sup>] | 7 (12.3) | 1.1 [0.43; 1199] |
| **Cancer type** |  |  |  |  |  |  |  |  |
| solid | 49 (72.1) | 32.03 [1.0; 29110] | 46 (76.7) | 2.1 [0.6; 15.0] | 46 (80.7) | 26629.3 [233; 6041000] | 41 (71.9) | 107.1 [2.0; 99710] |
| haematological | 19 (27.9) | 16.28 [1.0; 22690] | 14 (23.3) | 1.0 [0.3; 4.7] | 11 (19.3) | 20495.0 [72.0; 4.0 *10<sup>8</sup>] | 16 (28.1) | 1069.38 [15.0; 1426000] |

Note: The Overall column describes the values across all three endpoints. This means that a single trial can be included multiple times here (once for each available outcome).

### Endpoints

Statistical evidence for a beneficial effect on at least one endpoint was moderate for five out of 68 (7.4%), and (very) strong for 49 out of 68 (86.8%) indications. The median *BF* for OS was 1.9 (IQR 0.5-14.5), for PFS 24,767.8 (IQR 109.0-7.3*10^6^) and for TR 113.9 (IQR 3.0 - 547,100).

### Active control groups

Trials with active control groups (*n* =22) had a median *BF* of 17.9 (IQR 1.0 - 12,220.0) across all three endpoints. Three (13.6%) indications lacked (very) strong statistical evidence for benefits on any endpoint, while five indications (22.7%) were supported by (very) strong statistical evidence for benefits across all three endpoints.

Of the 19 indications with data available for OS, two (10.5%) were approved with pro-null or ambiguous statistical evidence for OS improvements without (very) strong evidence for improvements on surrogate endpoints. Additionally, 12 indications (63.2%) were approved based on pro-null or ambiguous statistical evidence for OS improvements but with (very) strong statistical evidence for improvements on at least one surrogate outcome. One indication (5.3%) was approved based on moderate statistical evidence for OS improvements and (very) strong evidence for improvements on the surrogate outcomes. The remaining four (21.1%) indications were approved based on (very) strong statistical evidence for OS improvements.

For PFS or TR outcomes, the majority of trials indicated (very) strong statistical evidence for better treatment outcomes in the experimental compared to the control group (*n* = 15, 79.0% and *n* = 16, 76.2% respectively; see also Figure 3). One drug (i.e., dasatinib) was approved based on ambiguous statistical evidence for TR only.

**Figure 3.**
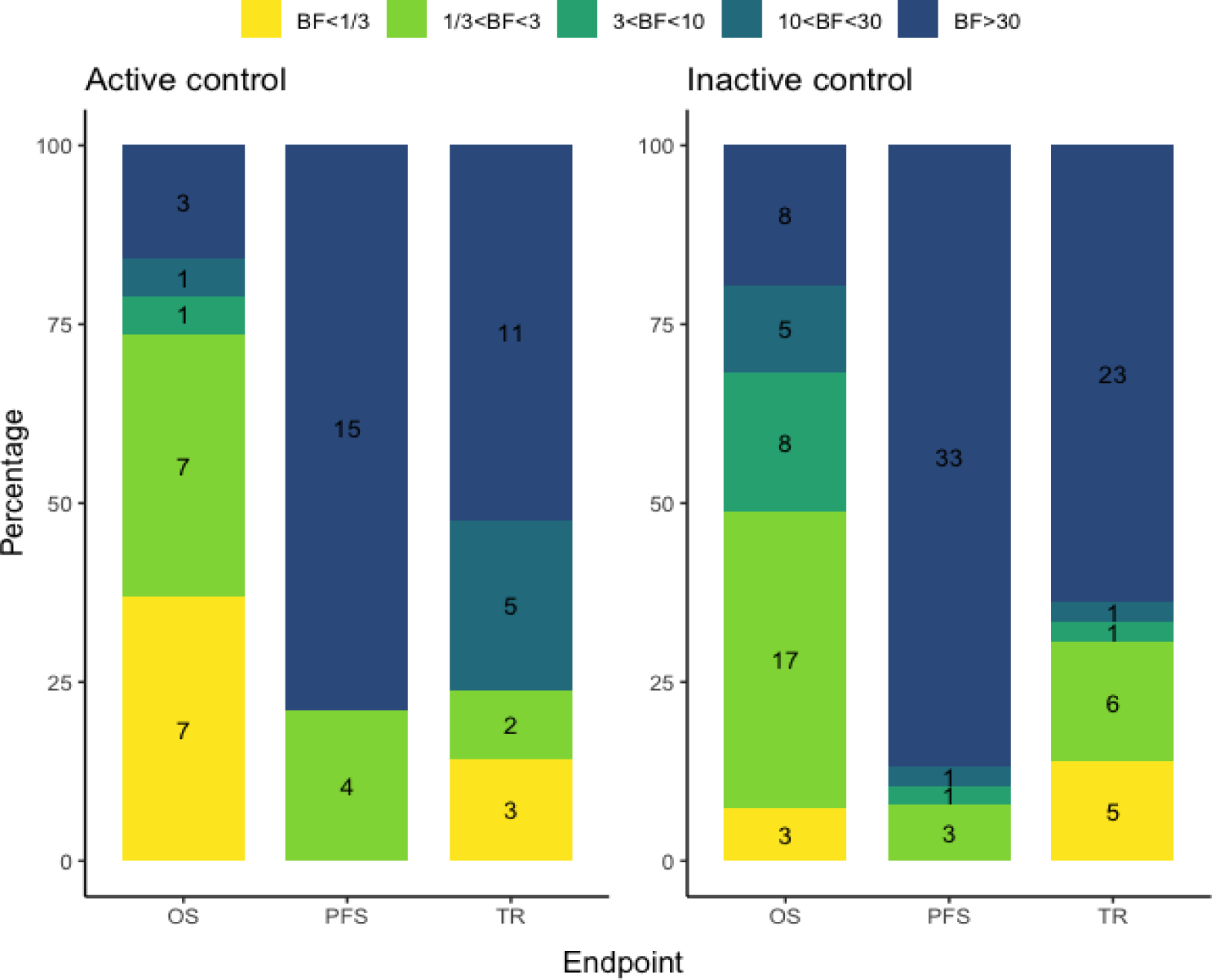
Bar plot illustrating the proportion of trials supported by pro-null evidence (BF<⅓), ambiguous evidence (⅓<BF<3), moderate pro-alternative evidence (3<BF<10), strong pro-alternative evidence (10<BF<30) and very strong pro-alternative evidence per endpoint and control condition. Counts are presented per category. Note that indications based on multiple trials are not included in this figure.

### Inactive control groups

Trials with inactive control groups (placebo: *n* = 29; supportive care: *17*) had a median *BF* of 58.2 (IQR 2 – 47,080.0) across all three endpoints. Two (4.3%) indications lacked (very) strong evidence for benefits on any endpoint (questioning the efficacy of the drug), while seven (17.1%) were supported by (very) strong statistical evidence for benefits across all three endpoints.

Of 41 indications with data available for OS, two (4.9%) were approved with pro-null or ambiguous statistical evidence for improvements of OS and all other endpoints. Additionally, 18 drugs (43.9%) were approved based on pro-null or ambiguous statistical evidence for OS improvements but with (very) strong statistical evidence for improvements on at least one surrogate outcome. Eight (19.5%) indications were approved based on moderate statistical evidence for OS improvements, of which five were supplemented with very strong statistical evidence for improvements of at least one surrogate endpoint. The remaining 13 (31.7%) indications were supported by (very) strong statistical evidence for OS.

The majority of indications were supported by very strong statistical evidence for improvements of PFS or TR (*n* = 34, 89.5% and *n* = 24, 66.7% respectively; see also Figure 3). One drug (neratinib maleate) was approved based on moderate statistical evidence for PFS improvements.

### Accelerated approval

Strength of statistical evidence was consistently lower for accelerated approvals (*n =* 7) across endpoints (Figure 4). No indications with accelerated approval lacked or provided (very) strong statistical evidence for improvements across all three endpoints. For indications that received non-accelerated approval, 5 of 58 indications lacked strong evidence for improvements on any endpoint, while 12 (out of 58) had strong or very strong statistical evidence for improvements across all three endpoints.

**Figure 4.**
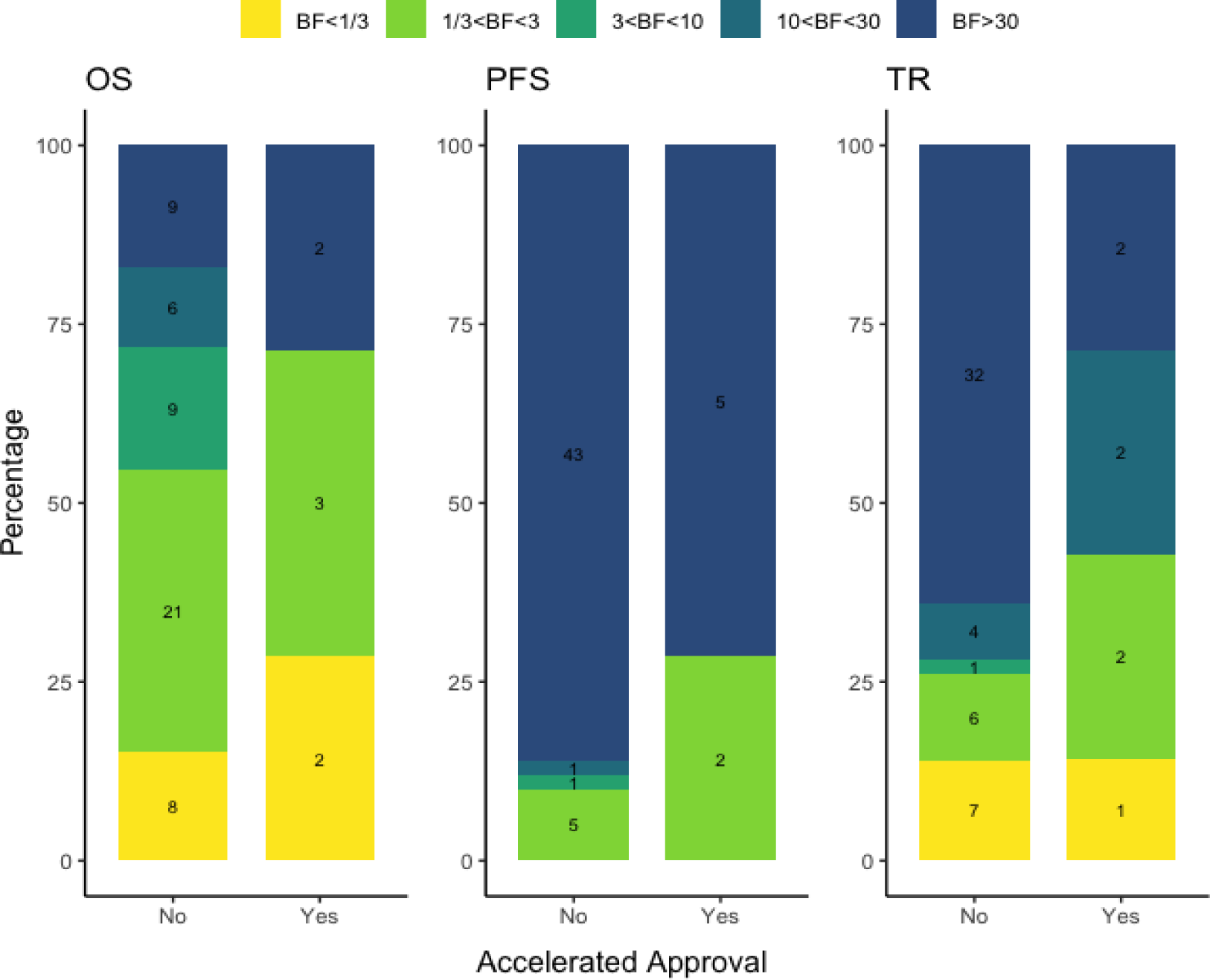
Bar plot illustrating the proportion of trials supported by pro-null evidence (BF<⅓), ambiguous evidence (⅓<BF<3), moderate pro-alternative evidence (3<BF<10), strong pro-alternative evidence (10<BF<30) and very strong pro-alternative evidence per endpoint and approval pathway. Note that indications based on multiple trials are not included in this figure.

Five out of seven (71.4%) accelerated approval decisions were based on pro-null or ambiguous statistical evidence for OS improvements, compared to 29 out of 53 (54.7%) non-accelerated approval decisions. Two (20.0%) accelerated approvals were based on pro-null evidence for OS improvements (active control: nivolumab; placebo control: panitumumab).

For PFS, no trial provided pro-null statistical evidence. The proportion of ambiguous statistical evidence was higher for accelerated approvals (2/7=28.6%) compared to non-accelerated approval (5/50=10.0%). For TR, the proportion of pro-null evidence was similar across approval pathways (accelerated: 1/7=14.3%; non-accelerated: 7/50=14.0%), but accelerated approvals were more frequently (2/7=28.6%) based on ambiguous statistical evidence compared to non-accelerated approvals (6/50=12.0%).

### Line of treatment

For OS and PFS, strength of statistical evidence did not differ qualitatively between lines of treatment (see Table 1). Although median strength of statistical evidence was greater for first-line treatment than for second-line treatment, IQRs mostly overlapped. For TR, strength of statistical evidence was lowest for trials supporting drugs approved for third or later line of treatment.

### Cancer type

There was no difference in strength of statistical evidence between solid and haematological cancer types (see Table 1).

### Approvals based on two RCTs

Seven drugs were approved based on two RCTs (Table 2).

**Table 2.** BFs corresponding to the individual trials and the meta-analytic BF for all drugs with more than one RCT. BFs are presented per outcome. Yellow: BF<⅓; Light green: ⅓ <BF<3, Dark green: 3<BF<10, Turquoise: 10<BF<30; Dark blue: BF>30.

| Drug | Outcome | Strength of Statistical Evidence (BF) |  |  |
| --- | --- | --- | --- | --- |
|  |  | RCT 1 | RCT 2 | Combined |
| fulvestrant | OS | 0.11 | 0.08 | 0.07 |
|  | PFS | - | - | - |
|  | TR | 0.28 | 0.09 | 0.33 |
| ruxolitinib phosphate | OS | 0.95 | 0.81 | 0.89 |
|  | PFS | 0.53 | - | - |
|  | TR | - | - | - |
| trastuzumab emtansine* | OS | 0.34 | 77.48 | 40.26 |
|  | PFS | 52279.93 | - | - |
|  | TR | 0.26 | 78.61 | 76.30 |
| bevacizumab | OS | 2.74 | 768.15 | 2060.87 |
| | PFS | 20.34 | $7.81 \times 10^8$ | $1.38 \times 10^9$ |
|  | TR | 2.70 | 5.89 | 25.6 |
| trifluridine tipiracil | OS | 30.20 | 1865.24 | 29179.5 |
| | PFS | $1.79 \times 10^{15}$ | - | - |
|  | TR | 0.04 | 0.05 | 0.71 |
| binimetinib | OS | - | 79.94 | - |
|  | PFS | 2.62 | 1677.46 | 4286.21 |
|  | TR | 4.49 | 2.11 | 45.4 |
| sorafenib tosylate | OS | - | 4.48 | - |
| | PFS | 697.30 | $5.96 \times 10^{10}$ | $1.20 \times 10^{10}$ |
|  | TR | - | 0.90 | - |

\*Please note that for this indication the participant groups differed between the RCTs; RCT1 was conducted in previously treated patients and RCT2 in previously untreated patients.

Two drugs (28.6%) were approved despite both trials providing pro-null or ambiguous statistical evidence. Of these, fulvestrant was approved based on two RCTs indicating that the treatment did not perform better than the active control on OS (meta *BF* = 0.07) and TR (meta *BF* = 0.12). The other, ruxolitinib, was approved based on two RCTs providing ambiguous statistical evidence for OS (meta *BF* = 0.89) and ambiguous statistical evidence for PFS from one trial (*BF* = 0.53). For the other five drugs, one RCT with ambiguous statistical evidence was supplemented by another RCT with at least moderate statistical evidence in favour of a treatment effect.

### Exploratory analysis

Relationship between strength of statistical evidence, effect size, and sample size In most cases, BF and 95% confidence intervals (CIs) were in agreement that an effect was pro-alternative (n=89) or ambiguous (n=28; i.e., 95% CI included the null). However, in 12 cases the 95% CI indicated uncertainty, while the *BF* was more informative, indicating absence of efficacy (lower left and right quadrant in Figure 5). Additionally, there were 12 effects for which the HR and corresponding confidence intervals indicated efficacy, whereas the *BF* indicated ambiguous statistical evidence.

**Figure 5.**
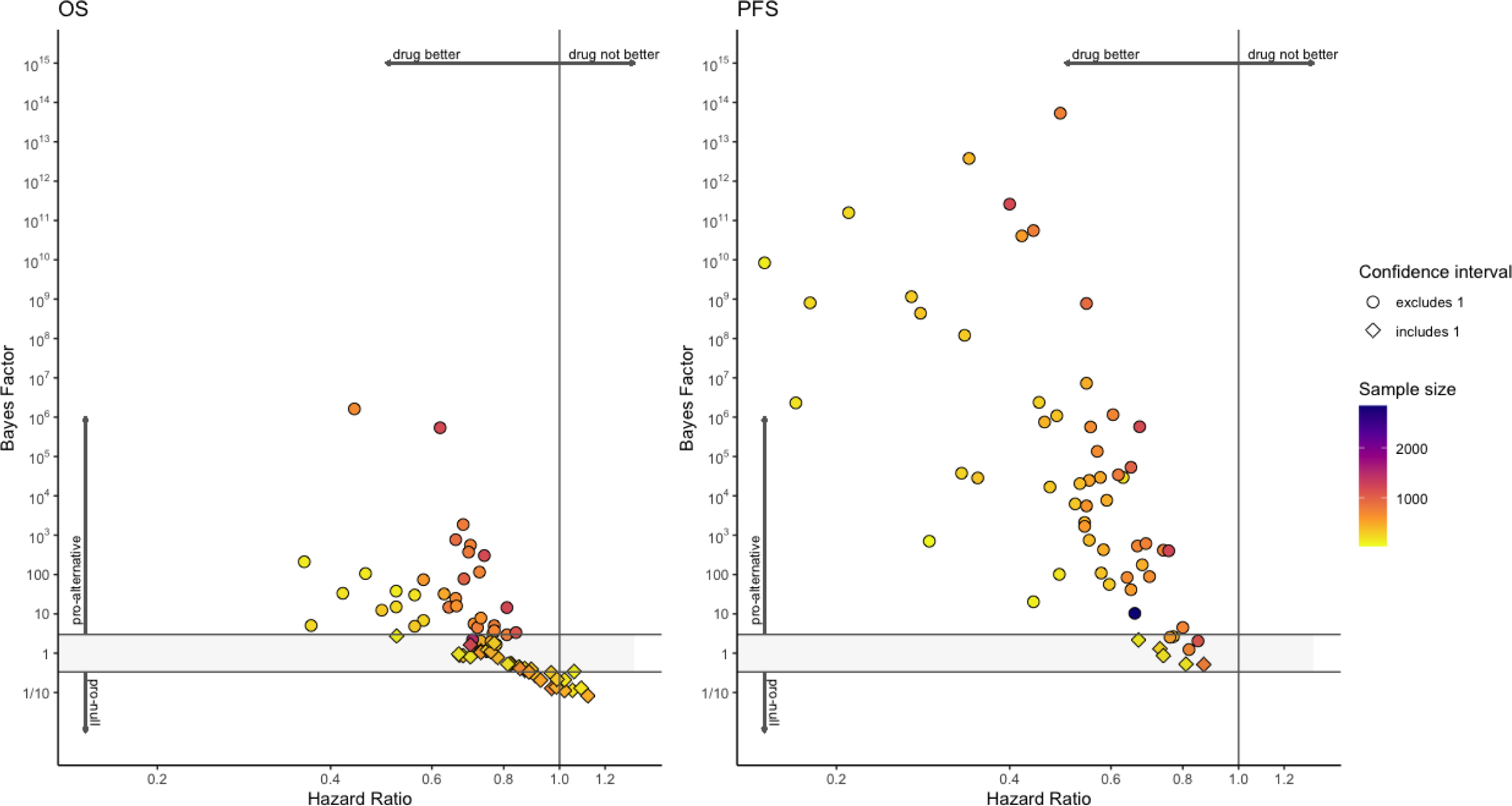
BFs plotted against HR for the 68 indications with one RCT. Shapes indicate whether or not the confidence interval of the HR included.

There was no relationship between strength of statistical evidence and sample size (*r* = - 0.03).

Results from the exploratory qualitative analysis are presented in Table 3 and detailed in the supplement (section 3).

**Table 3.** Reasons for drug approval mentioned for the seven drugs without statistical evidence for OS, PFS, or TR improvements. These themes were extracted via an exploratory qualitative thematic analysis.

| Theme | Subthemes |
| --- | --- |
| **Expected benefits over (non)-existing treatments due to...** | - poor prognosis (despite available treatments)<br>- few or highly toxic available treatments<br>- prior failed treatments<br>- different administration (e.g., oral medication) |
| **Tolerability** | - limited crossover between treatment arms<br>- low discontinuation rates |
| **Positive interpretation of limited statistical evidence** | - meaningful clinical differences (e.g., differences being statistically not significant but effect size deemed clinically meaningful)<br>- convincing interim analyses (e.g., very small *p*-value interpreted as robust statistical evidence)<br>- descriptive interpretation of premature survival analysis (e.g., eyeballing Kaplan-Meier survival curves or comparing percentages)<br>- non-inferiority to control group (e.g., statistically non-significant superiority trial interpreted as successful non-inferiority trial) |
| **Primary endpoints different from OS, PFS, or TR** | - regulatory precedents (e.g., other drugs in the same group were approved based on this endpoint)<br>- experience (e.g., the alternative endpoint was known to relate to OS or PFS) |
| **Use of non-randomized evidence** | - control group deemed unethical |

### Strength of statistical strength for primary endpoints

The median strength of statistical evidence was greater for OS and PFS when these outcomes were considered primary, compared to when they were not, although the interquartile IQRs mostly overlapped (see Table 4). Additional exploration regarding the differences in evidential strength between outcomes depending on which outcome was considered primary are presented in the supplement.

**Table 4.**
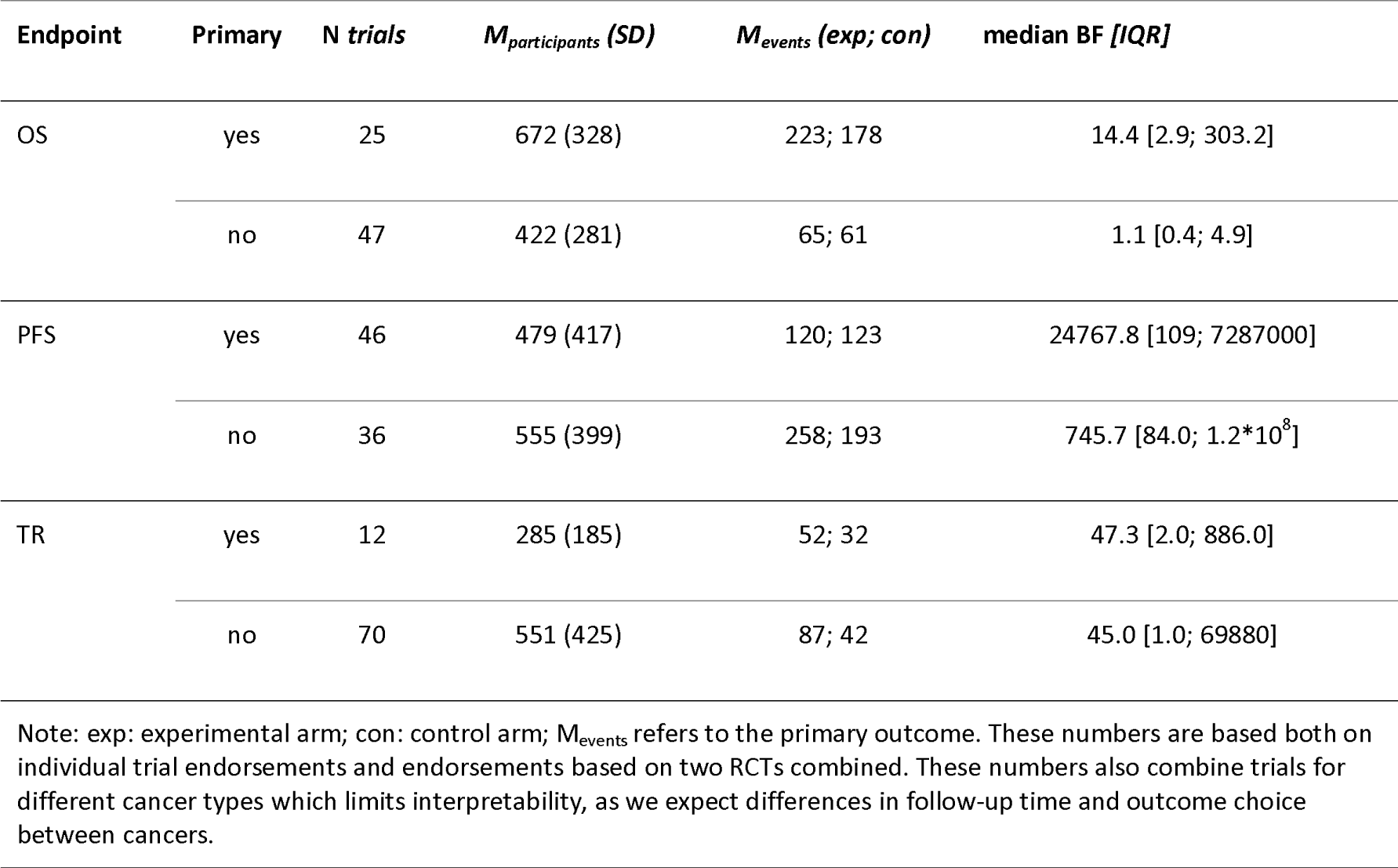
Median strength of statistical evidence for the primary and non-primary endpoints.

| Endpoint | Primary | N trials | $M_{participants}$ (SD) | $M_{events}$ (exp; con) | median BF [IQR] |
| --- | --- | --- | --- | --- | --- |
| OS | yes | 25 | 672 (328) | 223; 178 | 14.4 [2.9; 303.2] |
|  | no | 47 | 422 (281) | 65; 61 | 1.1 [0.4; 4.9] |
| PFS | yes | 46 | 479 (417) | 120; 123 | 24767.8 [109; 7287000] |
|  | no | 36 | 555 (399) | 258; 193 | 745.7 [84.0; 1.2*10<sup>8</sup>] |
| TR | yes | 12 | 285 (185) | 52; 32 | 47.3 [2.0; 886.0] |
|  | no | 70 | 551 (425) | 87; 42 | 45.0 [1.0; 69880] |

Note: exp: experimental arm; con: control arm; $M_{events}$ refers to the primary outcome. These numbers are based both on individual trial endorsements and endorsements based on two RCTs combined. These numbers also combine trials for different cancer types which limits interpretability, as we expect differences in follow-up time and outcome choice between cancers.

## DISCUSSION

Quantifying the strength of statistical evidence for efficacy for pivotal RCTs supporting novel cancer drugs approved by the FDA in the last two decades, we were able to provide explicit evidence that strength of statistical evidence was substantially lower for OS, arguably the most important outcome for cancer patients, compared to surrogate outcomes. Most indications (58.7%, 44/75) were approved without clear statistical evidence for OS improvements. While most of these indications (33/44) were supplemented by strong statistical evidence for improvements on at least one surrogate endpoint (i.e., PFS/TR), uncertainties regarding OS improvements remain. The present analysis using the *BF* was more informative than traditional measures of uncertainty such as the confidence interval, because it allowed us to disambiguate between absence of evidence and evidence of absence.

Strength of statistical evidence for efficacy differed between approval pathways but not lines of treatment or cancer types. Although few indications were approved through accelerated approval, our results suggest that weaker statistical evidence is accepted for accelerated approval decisions. While the FDA accepts higher levels of uncertainty for accelerated approval decisions due to the use of surrogate endpoints or intermediate clinical endpoints [25], we provide quantification of how much uncertainty the FDA considers acceptable. This again highlights the importance of timely post-approval studies to confirm efficacy. This need is also recognized in the Consolidated Appropriations Act, 2023 (H.R. 2617), enabling the FDA to require post-approval studies to be “*underway prior to granting accelerated approval*”[26]. It remains unclear how consistently the FDA will react to post-approval trials failing to confirm clinical benefits. As of 2021, one third of the novel cancer drugs receiving accelerated approval until 2020 but subsequently failing to improve primary endpoints in their post-approval studies, remained on the drug’s labelling (i.e., approved under the accelerated pathway) or were converted to regular approval [27]. Clear regulations and consistent action in response to post-approval trials is still lacking [27].

We observed indications receiving non-accelerated approval with absence of statistical evidence or even statistical evidence for the absence of efficacy based on other considerations. For example, favourable benefit-harm assessments were justified by expected benefits such as improved quality of life and safety profiles. However, reporting of quality-of-life-outcomes is incomplete [7] and not systematic, and a good safety profile is irrelevant in the absence of efficacy. In some cases, efficacy was determined in a manner different to the pre-registered protocol, for instance through eyeballing Kaplan-Meier survival curves or comparing percentages. We observed one instance in which a trial that failed to demonstrate superiority was re-interpreted as a non-inferiority trial. Switching between superiority and non-inferiority interpretations after results are known is problematic [28,29]. It would be preferable to supplement approval decisions with additional trials, which would lead to stronger statistical evidence.

In this paper, we primarily focused on cases with weak statistical evidence. However, there was significant variation in the strength of evidence, with *BF*s for PFS and TR often suggesting solid evidence for treatment effects. One might wonder, especially when time is of the essence, as it is with cancer drugs, whether these drugs could be approved on the basis of weaker evidence.

However, the discussion of how much evidence is needed to determine efficacy based on surrogate endpoints goes beyond the scope of this descriptive project, especially since this question cannot be separated from the question of how well (*BFs* of) surrogate outcomes predict (*BFs* of) OS. These questions warrant further investigation in future work.

### Strength and Limitations

We used a comprehensive database of pre-approval trials, examined multiple endpoints, and used a Bayesian approach to gain novel insights into the strength of evidence that allows for interpretations that are in line with clinical decision making and may provide a more intuitive perspective on evidence. The study also has several limitations. First, we focused on the statistical evidence of RCTs supporting approval decisions. This does not reflect the full complexity of the approval process and the variety of sources of evidence (e.g., quality of life, fewer side effects, method of administration etc.) that might be considered. Nevertheless, RCTs generally provide the strongest available evidence, as other sources of evidence (such as single-trial arms) are difficult to interpret. Second, interpretation of the *BF* depends on the control group. While we differentiate between active and inactive-controlled trials we did not classify whether comparators met the standard of care. As a result, some of our *BF* might be an overestimation of the strength of evidence for efficacy. Third, strength of statistical evidence is only one component of benefit assessment, and other factors also play a role, including the effect size and risk of bias. Fourth, we restricted our analyses to default priors to ensure comparability of our results across drug groups. Informed priors might be used in future analysis of individual drugs to integrate the available evidence into the statistical analysis. Lastly, we only included trials conducted to support approval decisions and did not include, for instance, post-approval studies. Therefore, our results indicate the strength of statistical evidence available to support initial approval decisions and do not necessarily reflect the current strength of statistical evidence.

## Conclusion

Regulatory decision making could be improved by using *BF*s to distinguish between drugs with good statistical evidence, drugs that lack statistical evidence, and drugs with statistical evidence against efficacy. We found that across the board the level of evidence for beneficial effects on OS is low. Average strength of statistical evidence on OS was moderate only if OS was considered the primary endpoint. While this suggests that evidence is better if the endpoint is considered primary, OS should be important regardless of whether it is a primary outcome or not. Some drugs were even approved without supporting statistical evidence on either OS, PFS or TR. These cases require a transparent and clear explanation. In many cases the statistical evidence is ambiguous, calling for additional trials, before or after approval, to reduce this uncertainty.

### List of Abbreviations

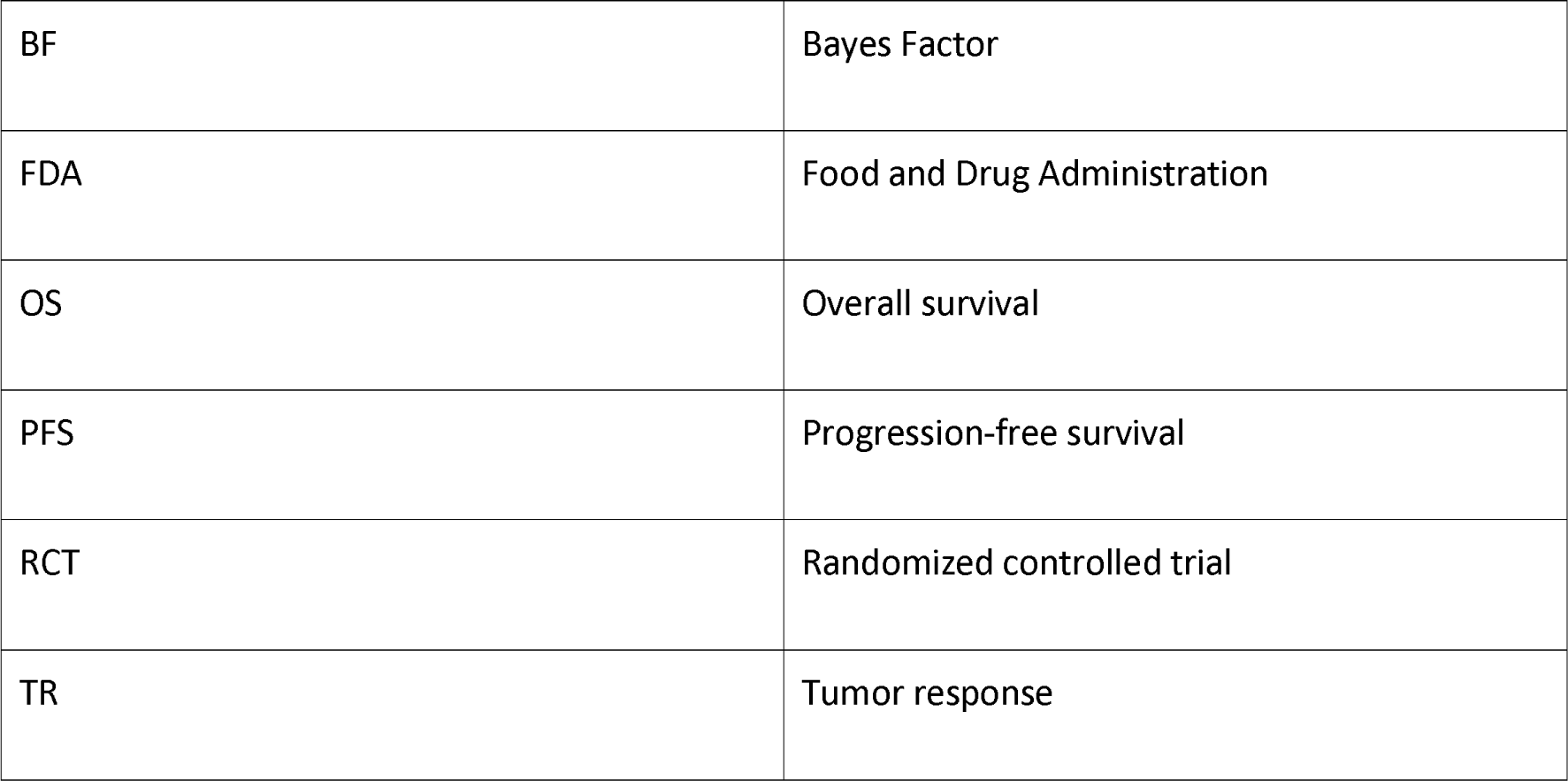

## Declarations

### Ethics approval

This study involved publicly available trial-level data. No ethical approval was needed.

## Consent for publication

Not applicable.

## Availability of data and materials

The data that support the findings of this study are available from https://ceit-cancer.org/ and the OSF framework (https://osf.io/4uhz7) DOI 10.17605/OSF.IO/QZ7XY.

## Authors’ contributions

***M.-M.P***.: Conceptualization, Data curation, Formal analysis, Methodology, Project administration, Resources, Visualization, and Writing - original draft. ***M.L***.: Formal analysis, Software, and Writing - review & editing. ***Y.A.d.V.***: Conceptualization, Supervision, and Writing - review & editing. ***L.G.H***.: Conceptualization, Investigation, and Writing - review & editing. ***A.M.S***.: Investigation and Writing - review & editing. ***R.R.M.***: Conceptualization, Supervision, and Writing - review & editing. ***D.v.R***.: Conceptualization, Funding acquisition, Supervision, and Writing - review & editing.

## Patient and Public Involvement

Patients or the public were not involved in the design, or conduct, or reporting, or dissemination plans of our research.

## Data Availability

The data that support the findings of this study are available from https://ceit-cancer.org/ and the OSF framework (https://osf.io/4uhz7).

## Acknowledgements

We would like to thank Jonathan Kimmelman for his insightful and constructive comments during the review process.

## Footnotes

1 Novel meaning first FDA approvals as opposed to label extensions or new indications of previously approved drugs.

2 Note that for ruxolitinib phosphate evidence was pooled across trials with active and inactive comparators.

3 Please note that we are specifically referring to instances in which there is statistical evidence for the absence of efficacy. We do not mean instances in which there is no evidence that the novel treatment is better than standard care (i.e., a pro-null or ambiguous BF for active controlled trials).

## References

1 Tafuri G, Stolk P, Trotta F, et al. How do the EMA and FDA decide which anticancer drugs make it to the market? A comparative qualitative study on decision makers’ views. Ann Oncol. 2014;25:265–9.

2 Ladanie A, Schmitt AM, Speich B, et al. Clinical Trial Evidence Supporting US Food and Drug Administration Approval of Novel Cancer Therapies Between 2000 and 2016. JAMA Netw Open. 2020;3:e2024406.

3 Gyawali B, Hey SP, Kesselheim AS. Assessment of the Clinical Benefit of Cancer Drugs Receiving Accelerated Approval. JAMA Intern Med. 2019;179:906.

4 Salas-Vega S, Iliopoulos O, Mossialos E. Assessment of Overall Survival, Quality of Life, and Safety Benefits Associated With New Cancer Medicines. JAMA Oncol. 2017;3:382–90.

5 Vokinger KN, Kesselheim AS. Characteristics of trials and regulatory pathways leading to US approval of innovative vs. non-innovative oncology drugs. Health Policy. 2019;123:721–7.

6 Michaeli DT, Michaeli T. Overall Survival, Progression-Free Survival, and Tumor Response Benefit Supporting Initial US Food and Drug Administration Approval and Indication Extension of New Cancer Drugs, 2003-2021. J Clin Oncol Off J Am Soc Clin Oncol. 2022;JCO2200535.

7 Gloy V, Schmitt AM, Düblin P, et al. The Evidence Base of US Food and Drug Administration Approvals of Novel Cancer Therapies from 2000 to 2020. Int J Cancer. 2023;152:2474–84.

8 U.S. Food and Drug Administration. Guidance for Industry: E9 Statistical Principles for Clinical Trials. 1998. https://www.fda.gov/media/71336/download

9 U.S. Food and Drug Administration. Qualification Process for Drug Development Tools. 2020.

10 Kemp R, Prasad V. Surrogate endpoints in oncology: when are they acceptable for regulatory and clinical decisions, and are they currently overused? BMC Med. 2017;15:1–7.

11 Pignatti F, Jonsson B, Blumenthal G, et al. Assessment of benefits and risks in development of targeted therapies for cancer — The view of regulatory authorities. Mol Oncol. 2015;9:1034–41.

12 Prasad V, Kim C, Burotto M, et al. The Strength of Association Between Surrogate End Points and Survival in Oncology: A Systematic Review of Trial-Level Meta-analyses. JAMA Intern Med. 2015;175:1389–98.

13 Gronau QF, Ly A, Wagenmakers E-J. Informed Bayesian T-Tests. Am Stat. 2019;1–14.

14 Rouder JN, Speckman PL, Sun D, et al. Bayesian t tests for accepting and rejecting the null hypothesis. Psychon Bull Rev. 2009;16:225–37.

15 Jeffreys H. Theory of probability. Oxford: Oxford University Press 1961.

16 Ladanie A, Speich B, Naudet F, et al. The Comparative Effectiveness of Innovative Treatments for Cancer (CEIT-Cancer) project: Rationale and design of the database and the collection of evidence available at approval of novel drugs. Trials. 2018;19:1–13.

17 Ladanie A, Ewald H, Kasenda B, et al. How to use FDA drug approval documents for evidence syntheses. BMJ. 2018;362:k2815.

18 R Core Team. R: A Language and Environment for Statistical Computing. Vienna, Austria: R Foundation for Statistical Computing 2021. https://www.R-project.org/

19 Linde M, van Ravenzwaaij D, Tendeiro JN. baymedr: Computation of Bayes Factors for Common Biomedical Designs. 2022. https://github.com/maxlinde/baymedr

20 Morey RD, Rouder JN, Jamil T, et al. BayesFactor: Computation of Bayes Factors for Common Designs. 2022. https://CRAN.R-project.org/package=BayesFactor (accessed 19 July 2022)

21 Jamil T, Ly A, Morey RD, et al. Default “Gunel and Dickey” Bayes factors for contingency tables. Behav Res Methods. 2017;49:638–52.

22 Heck DW, Gronau QF, Wagenmakers E-J. metaBMA: Bayesian model averaging for random and fixed effects meta-analysis. Retrieved Doi. 2017. https://cran.r-project.org/web/packages/metaBMA/metaBMA.pdf (accessed 20 March 2021)

23 Gronau QF, Heck DW, Berkhout SW, et al. A Primer on Bayesian Model-Averaged Meta-Analysis. Adv Methods Pract Psychol Sci. 2021;4:25152459211031256.

24 Lee MD, Wagenmakers E-J. Bayesian cognitive modeling: A practical course. Cambridge university press 2014.

25 U.S. Food and Drug Administration. Accelerated Approval. FDA. 2023. https://www.fda.gov/patients/fast-track-breakthrough-therapy-accelerated-approval-priority-review/accelerated-approval (accessed 30 January 2024)

26 New FDA Reform Legislation: Congress Gifts a “FDORA” for the Holidays. 2023. https://www.ropesgray.com/en/newsroom/alerts/2023/01/new-fda-reform-legislation-congress-gifts-a-fdora-for-the-holidays (accessed 16 February 2023)

27 Gyawali B, Rome BN, Kesselheim AS. Regulatory and clinical consequences of negative confirmatory trials of accelerated approval cancer drugs: retrospective observational study. BMJ. 2021;374:n1959.

28 U.S. Food and Drug Administration, Center for Drug Evaluation and Research. Non-Inferiority Clinical Trials to Establish Effectiveness Guidance for Industry. 2016. https://www.fda.gov/media/78504/download

29 Committee for Proprietary Medicinal Products. Points to consider on switching between superiority and non-inferiority. Br J Clin Pharmacol. 2001;52:223–8.

